# Isolation of extracellular vesicles from pleural effusion of patients with lung cancer for downstream application in the clinical setting

**DOI:** 10.64898/2025.11.28.25341238

**Authors:** Miodrag Vukovic, Lidija Filipovic, Nina Petrovic, Andrej Zecevic, Maja Kosanovic, Miljana Tanic, Radmila Jankovic, Tatjana Stanojkovic, Milica Popovic, Aleksandra Korac, Sanja Stevanovic, Milena Cavic

**Affiliations:** Department of Experimental Oncology, Institute for Oncology and Radiology of Serbia, Belgrade, Serbia; Department of Biochemistry, Faculty of Chemistry, University of Belgrade, Belgrade, Serbia; Laboratory for Radiobiology and Molecular Genetics, Department of Health and Environment, “VINČA” Institute of Nuclear Sciences-National Institute of the Republic of Serbia, University of Belgrade, Belgrade, Serbia; Clinic for Pulmonology, University Clinical Center of Serbia, Belgrade, Serbia; Institute for the Application of Nuclear Energy, INEP, University of Belgrade, Belgrade, Serbia; Faculty of Biology, Center for Electron Microscopy, University of Belgrade, Belgrade, Serbia; Institute of Chemistry, Technology and Metallurgy, Department of Electrochemistry, University of Belgrade, Belgrade, Serbia

**Author notes:** Corresponding author: Milena Cavic Department of Experimental Oncology Institute for Oncology and Radiology of Serbia (IORS) Pasterova 14, 11000 Belgrade, Serbia.

**Keywords:** Carcinoma, Extracellular Vesicles, Non-Small-Cell Lung Cancer, Pleural Effusion

## Abstract

Malignant pleural effusion (MPE) is a common clinical manifestation of advanced non-small cell lung cancer (NSCLC) and represents a valuable source of tumor-derived components, including extracellular vesicles (EVs). The aim of this study was to compare three different EV enrichment methods with potential applicability in clinical practice: a commercial Norgen kit (NOR), immunoaffinity capture (IA), and ultracentrifugation (UC). Following EV characterization, IA demonstrated the best overall performance in terms of EV yield and purity, NOR showed intermediate efficiency, while UC was the least effective method. The IA method exhibited characteristics suitable for potential clinical implementation, whereas NOR and UC may require combination with additional enrichment approaches. Advancing pleural effusion–based liquid biopsy toward clinical application will depend on the development of robust, scalable, and user-friendly EV enrichment workflows, along with harmonized guidelines for sample collection, preprocessing, and data reporting. Establishing such standards will enhance reproducibility, enable meaningful cross-study comparisons, and accelerate the integration of EV-based biomarkers into precision oncology.

## Introduction

Lung cancer remains one of the most common and deadliest malignancies worldwide, representing the leading cause of tumor-related mortality ^1^. Approximately 85% of cases are classified as non-small cell lung cancer (NSCLC), with a reported 5-year survival rate of only 25% ^2^. At the time of diagnosis, the majority of patients present with advanced disease, often accompanied by distant metastases, which substantially contributes to the poor prognosis and high mortality rates ^3^. In contrast, patients with early-stage NSCLC, defined as localized or locally advanced resectable disease, may achieve survival rates of up to 75%, particularly in younger individuals diagnosed with stage I disease ^4^. However, disease relapse occurs in 20–50% of patients with stage I–IIIA NSCLC, reducing 5-year survival rates to below 30% ^2^. These data highlight the urgent need for effective strategies aimed at early detection and timely therapeutic intervention. Although currently available screening and diagnostic tools, such as endobronchial ultrasound-guided fine needle aspiration (EBUS-FNA), magnetic resonance imaging (MRI), low-dose computed tomography (LDCT), and histopathological evaluation, provide valuable clinical information, their application is frequently limited by invasiveness, cost, radiation exposure, or procedural risks associated with tissue biopsies ^5–7^ as well as various implementation issues especially in countries with limited resources ^8^.

Liquid biopsy has already been established as a less invasive alternative, enabling repeated and longitudinal sampling in the clinical setting ^9^. Beyond initial tumor detection, liquid biopsy can provide information on pathological classification, staging, mutational profiling, monitoring of disease progression, and evaluation of treatment response ^10^. While blood is the most frequently used source, other biological fluids, including cerebrospinal fluid, saliva, ascites, urine, tears, breast milk, and pleural effusion (PE), have also been explored ^11,12^.

PE represents a frequent clinical manifestation of different conditions, such as malignancy, infection, autoimmune disorders, trauma and pulmonary embolism. Based on etiology, it can be classified as benign (heart failure, parapneumonic infections, etc.) or malignant. In lung cancer, malignant pleural effusion (MPE) develops either through direct tumor invasion of the pleura, or through haematogenic/lymphogenic spreading of tumor cells ^13^. The presence of MPE typically signifies advanced disease, with a median overall survival of 3–12 months following diagnosis and a 5-year overall survival rate of only 3% ^14,15^.

Thoracentesis is a minimal invasive procedure for collecting the pleural fluid, either for diagnostic purposes or to relive symptoms, such as shortness of breath. Its sampling is not required as an additional diagnostic step but is rather obtained during usual symptom-relieving measures in the clinic. Cytological and biochemical analysis of MPE is a routine diagnostic approach. Unfortunately, low tumor cell abundance frequently results in delayed or inconclusive diagnoses ^16^. However, it is a valuable source for downstream molecular analyses. MPE is rich in tumor-derived components with potential clinical relevance, including circulating tumor DNA (ctDNA), microRNAs (miRNAs), long non-coding RNAs (lncRNAs), circulating tumor cells (CTCs) and tumor-educated cell types, such as platelets and macrophages. In addition, MPE contain extracellular vesicles (EVs) that serve as carriers of various tumor-associated biomolecules ^17^.

EVs have recently attracted significant attention as potential diagnostic and prognostic biomarkers ^18^. They are commonly categorized into exosomes, EVs of endosomal origin released by the exocytosis of multivesicular bodies (MVBs) and ectosomes, which are produced by plasma membrane budding ^19^. EVs play key roles in maintaining cellular homeostasis and mediating intercellular communication by transferring proteins, nucleic acids, and lipids. Importantly, they can also facilitate tumor progression by shuttling oncogenic proteins, non-coding RNAs, and microRNAs, thereby altering gene regulation in recipient cells at the post-transcriptional level ^20–23^. Consequently, EVs infer the state of the parental cells and as such have been increasingly investigated as minimally invasive biomarkers for cancer detection and monitoring.

Despite the potential of EVs derived from MPE as a source of tumor-associated biomarkers, there is currently no standardized approach for their enrichment ^24^. Previous studies investigating MPE-derived EVs have employed heterogeneous methodologies, each with specific advantages and limitations. While combining multiple enrichment strategies has been suggested to improve yield and purity, such approaches are costly, labor-intensive, and difficult to implement in routine clinical practice. Standardization of EV enrichment methods in clinical setting is necessary for clinical implementation of MPE-EV. The International Society for Extracellular Vesicles (ISEV) has employed initiatives^*^ to address this important issue through the formation of Scientific Reproducibility Intersociety Working Groups (such as ISEV-ELBS with the European society for liquid biopsy). Recommendations have been published for other biological fluids ^25,26^, but there is currently no recommendation for MPE.

Therefore, the present study was designed to compare three different methods for EV enrichment from MPE samples, including two techniques, immunoaffinity capture and NORGEN commercial kit, not previously applied to this sample type. The aim was to identify a single method that provides sufficient yield and purity of EVs suitable for downstream omics molecular analyses, while maintaining feasibility for potential clinical application after process standardization even in countries with limited healthcare resources.

## Methods

### Pleural effusion sample collection

MPE samples were obtained from five patients with advanced lung adenocarcinoma, stage IV (three men and two women, age range from 52 to 71) by thoracentesis for therapeutic purposes, at the University Clinical Center of Serbia, Clinic for Pulmonology. The samples were processed within one hour after thoracentesis. To remove cells and cellular debris, the samples were centrifuged for 10 minutes at 300 x g, followed by 15 minutes at 2000 x g. The samples were then frozen at -80°C. Before starting the isolation of EVs, the samples were thawed and centrifuged again for 10 minutes at 2000 x g to remove cryoprecipitate. For EVs isolation MPE pool was made from these five patients, and diluted with PBS (ratio 1:1). Prepared pooled sample was divided into aliquots, so that each isolation method could be performed in triplicate. All analyses from this study were approved by institutional Ethics Committees (approval no. 3780/2 from 07.11.2022. and 01-1/2024/965 from 29.3.2024.) and all patients signed an informed consent.

### Enrichment of EVs with three different methods: NORGEN commercial kit (NOR), Immunoaffinity capture (IA) and Ultracentrifugation (UC)

EVs enrichment using the NOR was carried out according to the manufacturer’s instructions (https://norgenbiotek.com/sites/default/files/resources/Plasma-Serum-Exosome-Purification-and-RNA-Isolation-Mini-Kit-Insert-PI58300-1.pdf?srsltid=AfmBOooqP8XTpWF83B_nUuxleoCixhqceqmRh5Y_hypfhHskA5yeGVDa). The complete protocol for EVs enrichment using the IA method is described in more detail in the publication by Filipovic et al ^27^. Briefly, 500 µL of methacrylate-based microporous copolymer, previously activated and functionalized with nanobodies targeting EVs, was incubated overnight at 4°C with 1 mL of MPE sample under continuous mixing. After incubation, the sample was removed, and the polymer with bound vesicles was washed several times with PBS. For the final elution step, 400 µL of 200 mM glycine buffer (pH 2.2) was added, followed by a 20-minute incubation. The eluate was then collected into a new tube and neutralized with 100 µL of 1 M Tris-HCl (pH 9.0). UC was performed according to the following protocol: 17,000 × g for 25 minutes and 100,000 × g for 2 hours on Optima L-90K ultracentrifuge, using SW 41 Ti rotor (Beckman Coulter, Indianapolis, IN, USA). Starting volume for MPE-EVs enrichment for IA and was 1mL of MPE pool, and 12 mL for UC.

### MPE-EVs Quantification

The size distribution and concentration of MPE-EVs were analyzed using the ZetaView® QUATT NTA system PMX-430 (Particle Metrix, Inning am Ammersee, Germany), operated with ZetaView software version 8.05.16 SP3. Instrument calibration, including camera and laser alignment and focus adjustment, was performed using 100 nm polystyrene beads, following the manufacturer’s guidelines. Samples were diluted in PBS to achieve an optimal particle concentration per frame. Measurements were conducted in scatter mode using a 488 nm blue laser. The camera was configured with a sensitivity of 78, shutter speed of 100, and a frame rate of 30 frames per second.

Post-acquisition analysis parameters were set as follows: minimum particle area of 10, maximum area of 200, and a minimum brightness threshold of 25. Raw data representing the number of particles detected at each vesicle size were exported and visualized using Microsoft Excel (Office 2016). Each analysis was performed in triplicate and results presented as mean values ± SD.

### Scanning electron microscopy

Scanning electron microscopy (SEM) was employed to examine the morphology of isolated MPE-EVs. The MPE-EVs samples were applied to carbon-coated grids and air-dried. Prior to SEM imaging, the samples were coated with a thin layer of gold using a sputter coater (Polaron SC503, Fisons Instruments, Glasgow, Scotland or Baltec SCD 005 sputtering chamber, New York, NY, USA). Morphological characterization of the MPE-EVs was performed using a Tescan Field Emission Scanning Electron Microscope (FE-SEM) Mira 3XMU (Tescan, Brno, Czech Republic). Image processing (cutting, merging, contrasting) was performed on Adobe Photoshop 2020 software.

### Atomic force microscopy

Surface morphology of MPE-EVs was analyzed using atomic force microscopy (AFM) on a NanoScope 3D system (Veeco, Plainview, NY, USA), operated in tapping mode under ambient conditions. Etched silicon probes with a spring constant ranging from 20 to 80 N/m were employed. Image analysis was conducted using Nanoscope image processing software (version 1.40r1). Prior to imaging, the mica substrate was mechanically polished using adhesive tape. A 10 µL aliquot of the EV suspension was deposited onto the polished mica surface and allowed to air dry.

### Transmission electron microscopy

The morphology of enriched MPE-EVs was assessed using transmission electron microscopy (TEM) following negative staining of the samples. EV suspensions were applied to carbon-coated grids, and after adsorption, excess liquid was removed. The EVs were then fixed with 2.5% glutaraldehyde and subsequently contrasted using 1% phosphotungstic acid. Samples were left to air dry prior to imaging. Electron micrographs were acquired using a Philips CM12 transmission electron microscope (Philips, Eindhoven, The Netherlands) equipped with an SIS MegaView III digital camera (Olympus Soft Imaging Solutions, Münster, Germany).

### Flow cytometry

Samples for flow cytometry analysis were prepared following previously described ^27^. In brief, 30 µL of a 1% bead suspension was incubated overnight at 4 °C with shaking, together with 4–40 µg of vesicle protein. The following day, beads were blocked first with 200 mM glycine for 30 minutes, followed by 5% skimmed milk in PBS. After blocking, beads were washed with PBS to remove residual blocking agents.

The bead suspension was then divided into three separate aliquots, each incubated for 1 hour with one of the following commercial antibodies: anti-CD9, anti-CD81, or anti-CD63 (each diluted 1:5, BioLegend, cat: 312106, 353038 and 349519, respectively). These three samples were analyzed individually using a BD FACSCalibur flow cytometer (BD Biosciences, Franklin Lakes, NJ, USA). Excitation was performed with a 488 nm blue laser, and fluorescence emissions were detected at the following wavelengths: anti-CD63 (AlexaFluor 488, FL1, 525 nm), anti-CD9 (PE, FL2, 561 nm), and anti-CD81 (PE/Dazzle, FL3, 620 nm).

Positive events were assessed relative to control beads (non-coated and blocked with milk). For negative control, a 1% Triton X-100 solution was added to each sample in a 1:1 ratio (resulting in a final concentration of 0.5%) and incubated for 45 minutes at room temperature. Samples were then reanalyzed under the same conditions.

### MPE-EVs protein and lipid concentration

The concentration of EV-associated proteins was determined using the Micro BCA Protein Assay Kit (Thermo Scientific™, catalog number 23235), with bovine serum albumin (BSA) employed as the standard. Lipid content in MPE-EVs was quantified using the colorimetric sulfophosphovanilin (SPV) assay.

## Data analysis

For data analysis, Microsoft Excel 2016 and GraphPad Prism v8 software were used for statistical calculations (mean and standard deviation) and one-way ANOVA with Tukey’s post hoc multiple comparison test. Flow cytometry data were analyzed using FlowJo X 10.0.7, and image processing was conducted with Microsoft PowerPoint 2016.

## Results

### NTA, protein and lipid concentration

The concentration and size distribution analysis of EVs obtained from MPE using three methods (NOR, IA, UC) was performed by NTA, yielding the following results: the mean values ± SD for vesicle diameter were 156 ± 5.5 nm for the NOR, 149 ± 0.3 nm for IA and 225 ± 7.6 nm for UC (Figure 1). The concentration of MPE-EVs was 4.29E+10, 1.15E+11 and 1.89E+10 particles/mL, respectively. Average protein concentration in MPE-EVs isolates was 1615 ± 12 µg/mL, 500 ± 87.6 µg/mL and 1260 ± 87 µg/mL, respectively. Average lipid concentration in MPE-EVs isolates was 116 ± 12 µg/mL, 374 ± 38 µg/mL and 13,6 ± 0.4 µg/mL, respectively (Table 1 and Figure 2). For EVs samples obtained by UC, the NTA particle concentration, as well as protein and lipid concentrations, were calculated by dividing the measured values by 12, since the initial volume used for EV isolation was 12 mL of pooled MPE.

**Figure 1.**
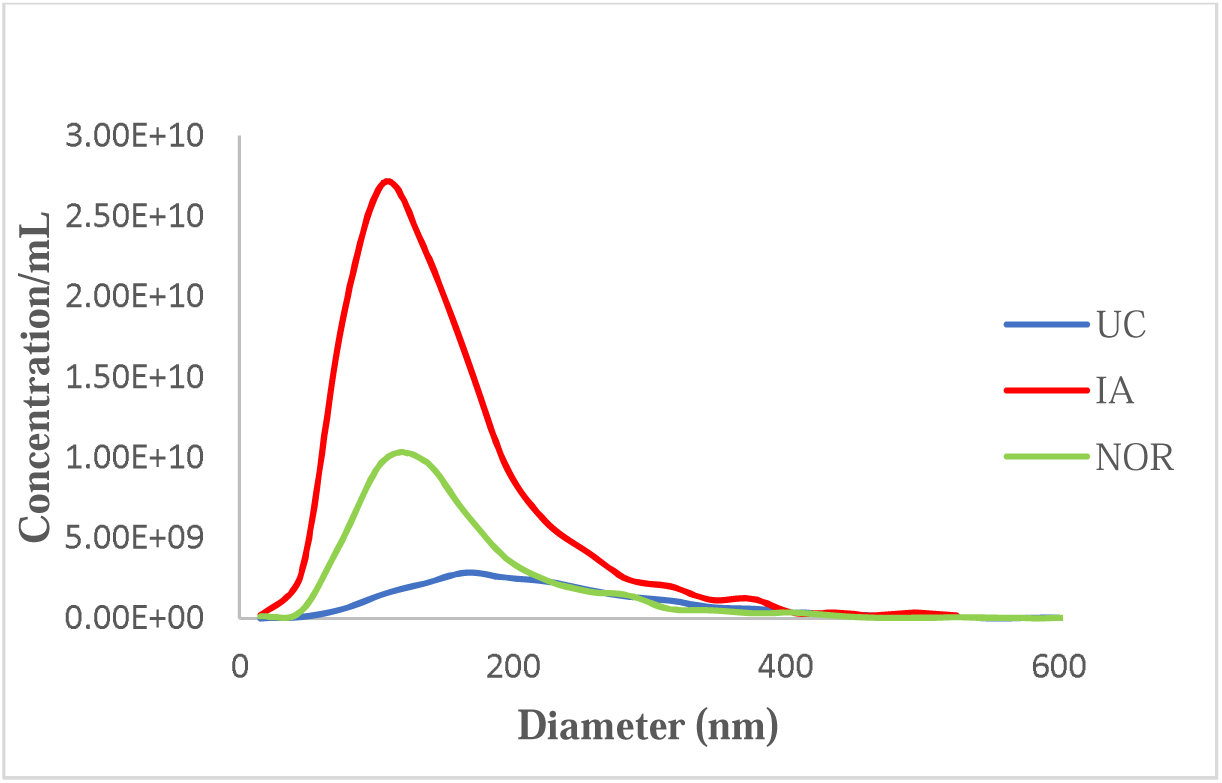
NTA results of MPE-EVs enriched with three methods

**Table 1.** Characteristics of isolated MPE-EVs

| Isolation | Volume of | Concentration | Average | Protein conc. | Lipid | Protein/Lipid |
| --- | --- | --- | --- | --- | --- | --- |

| method | MPE pool<br>(mL) | (particles/mL)<br>(summary) | size ± SD<br>(nm) | ($\mu\text{g/mL}$) ±<br>SD | conc.<br>($\mu\text{g/mL}$) ±<br>SD | ratio ± SD |
| --- | --- | --- | --- | --- | --- | --- |
| **NOR** | 1 | 4.29E+10 | 156.0±5.5 | 1615.0±149.6 | 116.0±12.0 | 12.8±0.9 |
| **IA** | 1 | 1.15E+11 | 149.0±0.3 | 500.0±87.6 | 374.0±38.0 | 1.3±0.2 |
| **UC** | 1 | 1.89E+10 | 225.0±7.6 | 1260.0±87.0 | 13.6±0.4 | 92.7±4.0 |

**Figure 2.**
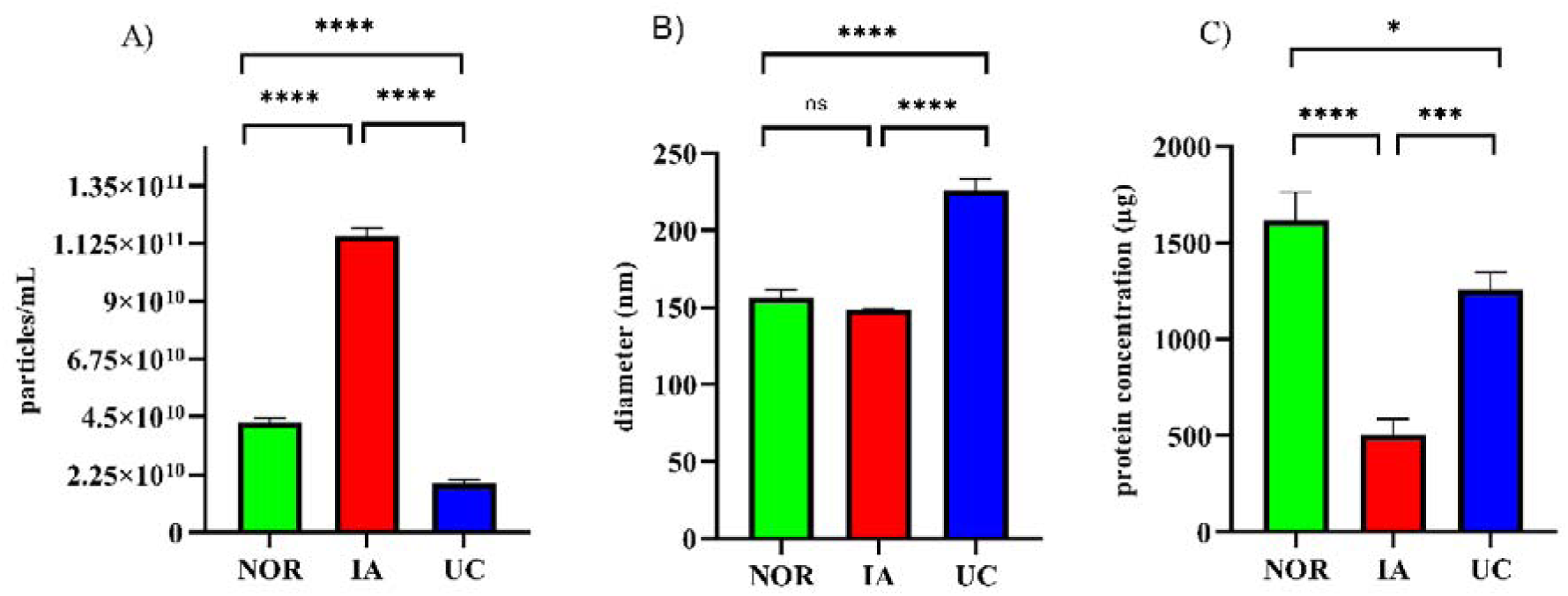
Comparison of MPE-EVs yield, size, and protein content obtained by three EVs enrichment methods. (A) Particle concentration (particles/mL), (B) mean particle diameter (nm), and (C) protein concentration (µg) was compared among NOR, IA, and UC methods using one-way ANOVA followed by Tukey’s multiple comparison post hoc test. Data are presented as mean ± SD. Statistical significance levels: **** p < 0.0001; *** p = 0.0008; * p = 0.03; ns – non significant.

### Flow cytometry

Flow cytometry was performed to assess the presence of surface antigens on MPE-EVs isolated using three different methods: NOR, IA, and UC. All methods demonstrated clear positivity for the established EV markers CD9 (33.15% for NOR, 29.95% for IA, 41.8% for UC), CD63 (38.7% for NOR, 26.50% for IA, 21.25% for UC), and CD81 (29.25% for NOR, 31.80% for IA, 32.35% for UC). Noticeable decrease in signal intensity was observed after treatment with 1% Triton X-100 for 45 minutes (reducing positivity of CD9, CD63 and CD81 for NOR, IA and UC to 21.35%, 18.65%, 37.20%; 33.90%, 16.30%, 15.05% and 17.60%, 20.85%, 25%, respectively)—confirming that the detected markers were associated with vesicular structures (Figure 3A, 3B, and 3C).

**Figure 3.**
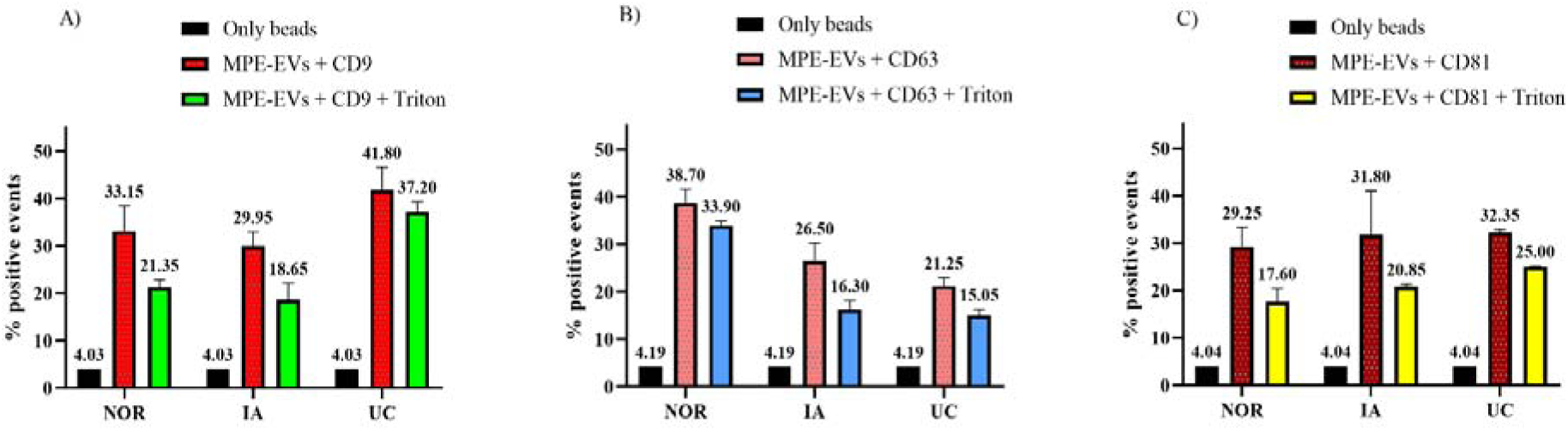
Flow cytometry analysis for CD9 (A), CD63 (B) and CD81 (C) positive MPE-EVs enriched with NOR, IA and UC before and after Triton X-100 treatment (mean ± SD).

### Scanning electron microscopy

The morphology of the isolated vesicles was examined using scanning electron microscopy (SEM). Both individual and clustered MPE-EVs were observed across the field of view, displaying a predominantly spherical shape (Figure 4). Their diameter was generally below 150 nm, consistent with the size range of small EVs. The morphology and degree of EVs aggregation varied depending on the isolation method, as observed by scanning electron microscopy (SEM). EVs isolated using the NOR (Figure 4A) appeared as well-dispersed, predominantly spherical particles with uniform size and clearly defined contours, without aggregation. Similarly, EVs isolated by IA (Figure 4B) also showed individual vesicles with minimal aggregation. On the other hand, EVs obtained through UC (Figure 4C) exhibited pronounced aggregation, forming dense clusters of vesicle-like structures with less clearly defined boundaries. These findings highlight the influence of the isolation method on EV morphology and physical state, which is particularly important for downstream analyses.

**Figure 4.**
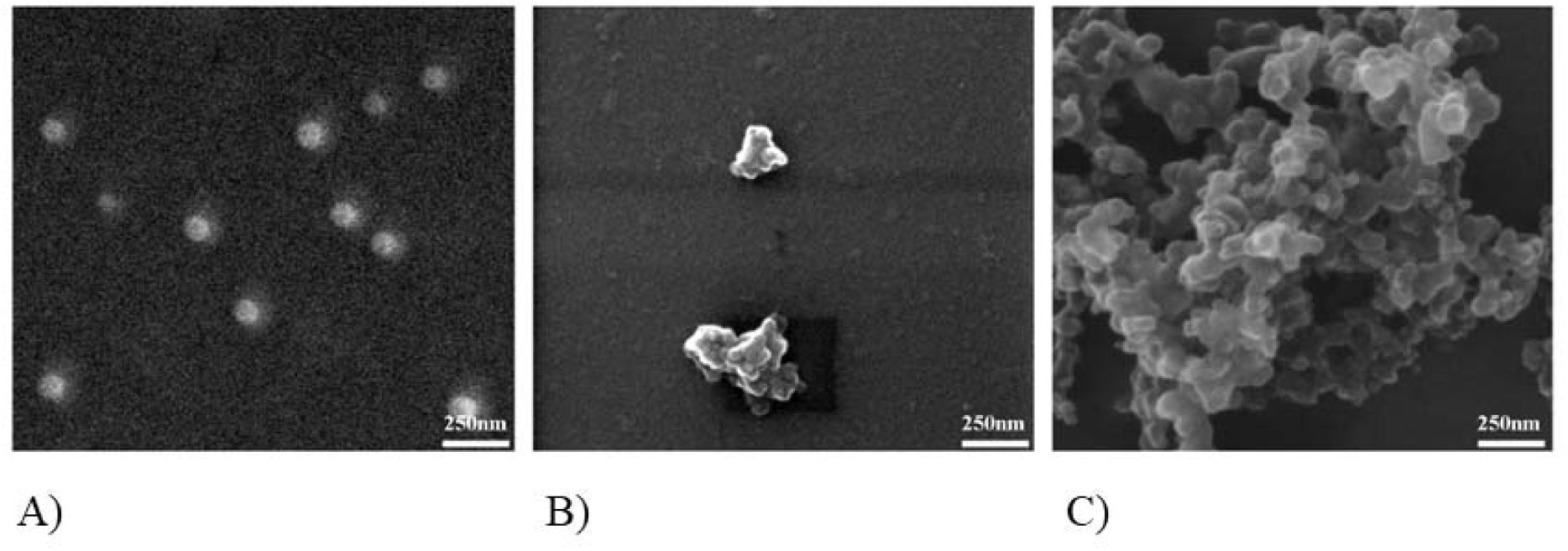
SEM analysis of MPE-EVs obtained with A) NOR, B) IA and C) UC

### Transmission electron microscopy

TEM analysis results showed that all three methods for EVs enrichment yielded similar outcomes regarding vesicle size. MPE-EVs obtained with all three methods exhibited diameters within the range of less than 200 nm, which is consistent with the known characteristics of small EVs (Figure 5).

**Figure 5.**
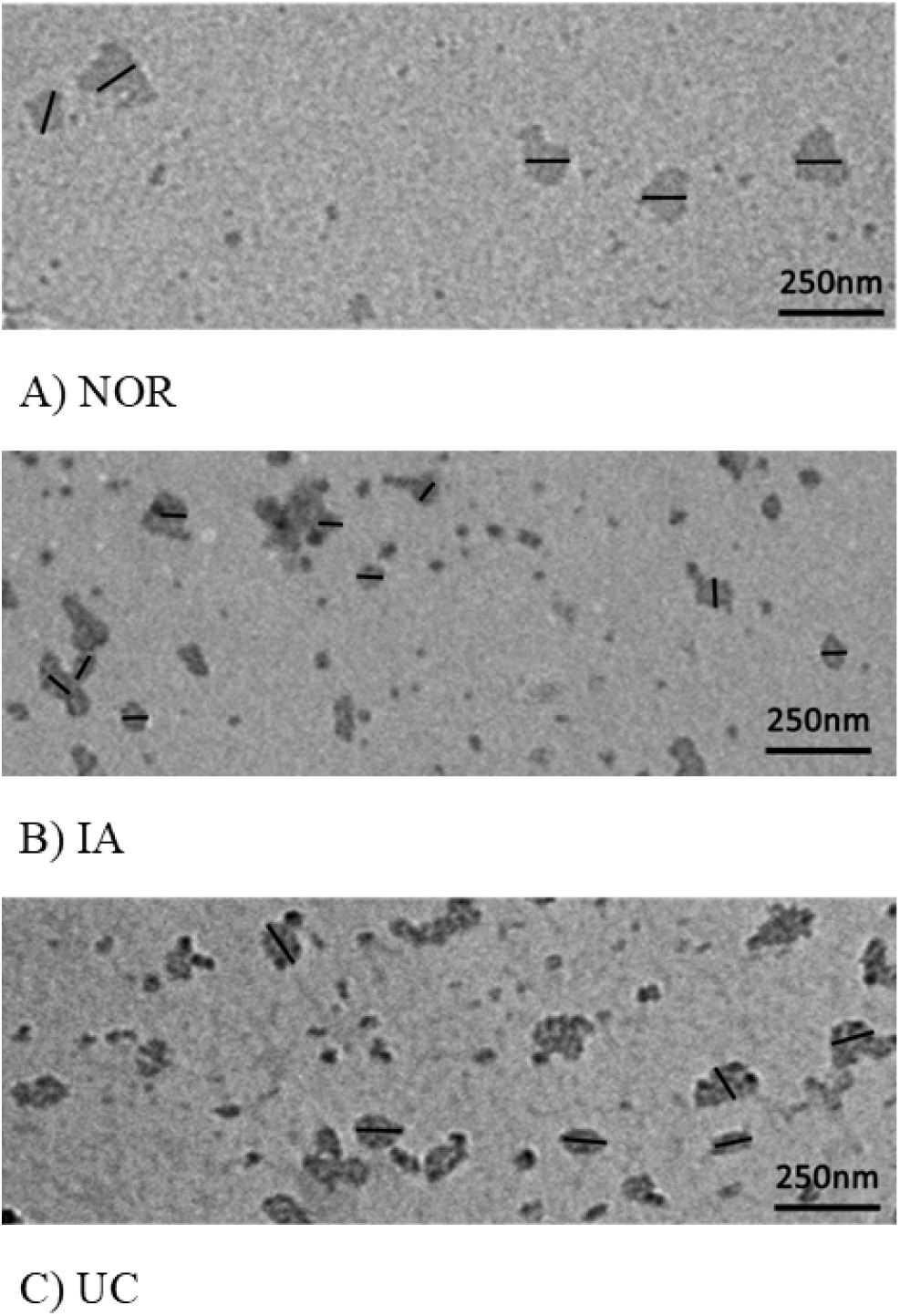
TEM analysis of MPE-EVs enriched with A) NOR, B) IA and C) UC

### AFM microscopy

Atomic force microscopy (AFM) was employed to further characterize the surface morphology of the MPE-EVs after their deposition on an atomically flat mica substrate (Figure 6). Representative two-dimensional (2D) and three-dimensional (3D) AFM images are shown in panels A, B and C, clearly revealing the spherical or globular appearance of the MPE-EVs. The size of individual vesicles was determined through cross-sectional profile analysis of the AFM images. The measured diameters varied depending on the isolation method: for MPE-EVs obtained using the NOR, diameters ranged from 84 to 88 nm, with red markers indicating vesicles of 85 nm, green 84 nm, and black 88 nm; for MPE-EVs obtained via IA, the vesicles were smaller, with diameters ranging from 30 to 49 nm. Red markers correspond to 49 nm vesicles, green to 45 nm, and black to 30 nm; for UC enriched MPE-EVs, vesicle diameters ranged from 47 to 85 nm, with red, green, and black markers denoting vesicles of 85 nm, 61 nm, and 47 nm in diameter, respectively. These findings highlight method-dependent differences in vesicle size and support the morphological integrity of the isolated MPE-EVs.

**Figure 6.**
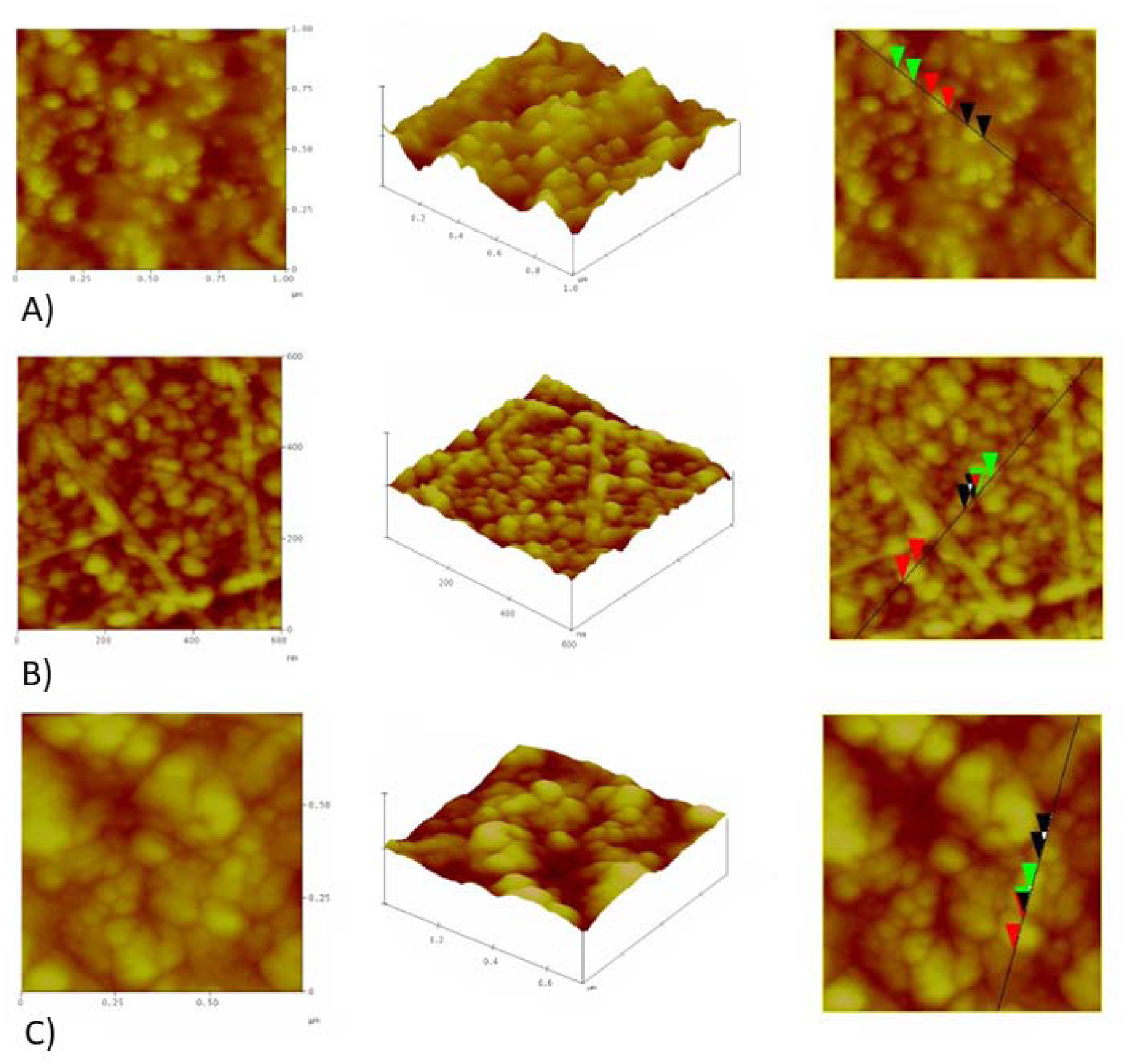
AFM characterization of MPE-EVs. Figures (A, B and C) show two-dimensional, three-dimensional topographical analyses and cross-sectional profile of the vesicle surface for NOR, IA and UC, respectively.

## Discussion

The current gold standard for differentiating benign from malignant PE is cytological examination of pleural fluid. However, cytology remains non-diagnostic in up to 70% of malignant effusions, necessitating more invasive biopsy procedures for definitive diagnosis ^28^. In addition to its low sensitivity, cytology does not provide information on the molecular profile of metastatic tumors, which is crucial for identifying patients eligible for personalized therapies. In this context, liquid biopsy applied to PE has emerged as a minimally invasive strategy capable of capturing tumor-derived biomarkers and potentially overcoming the limitations of conventional PE diagnostics ^17^.

The first characterization of small EVs in MPE was conducted by André et al., who described vesicles approximately 80 nm in diameter enriched in antigen-presenting molecules and tumor antigens ^29^. Since then, small EVs from MPE have been further characterized using proteomic approaches, although most studies have focused on non-coding RNAs, particularly miRNAs, as biomarkers to distinguish malignant from benign effusions ^30–32^. Despite these promising findings, no standardized method for isolating MPE-derived EVs has been established.

Ultracentrifugation (UC) has been the most widely used method in published studies. Nevertheless, UC is limited by its inability to efficiently separate EVs from abundant lipoproteins, such as HDL, which may interfere with downstream proteomic analyses ^33,34^. In our study, UC proved to be the least effective method, yielding low particle concentrations and larger particle diameters. Furthermore, EVs isolated by UC showed a tendency to aggregate, raising questions about whether the observed size distribution reflects a genuine abundance of larger vesicles or artefacts of aggregation, which is possible due to freeze-thaw cycles of EV isolates ^35^. Also, extensive aggregation is possible due to the high centrifugal forces applied during the isolation process, which can promote vesicle clustering and potentially compromise the structural integrity of individual particles.

By contrast, the IA method developed by Filipović et al. ^27^ at the University of Belgrade, Faculty of Chemistry demonstrated superior performance across multiple parameters, including concentration, diameter distribution, and purity of isolated vesicles. Although some degree of aggregation was also observed, it was less pronounced than with UC. Importantly, smaller vesicles were more consistently represented in their diameter across microscopy analyses. These results are consistent within all three methods for EV enrichment, especially in comparison with results from NTA, which is in accordance with literature data, where, for example, SEM and NTA significantly deviated for EVs smaller than 70nm ^36^. The NOR produced results intermediate between UC and IA in terms of vesicle concentration, where the particle concentration was the highest in IA. Its NTA profile closely resembled IA with respect to diameter distribution, though protein concentrations were comparable to UC. Interestingly, only NOR-derived EVs did not exhibit aggregation, which may represent a distinct advantage.

In addition to the conventional characterization approaches, the purity of MPE-EVs can also be assessed through the protein-to-lipid (P/L) ratio, which has been proposed as a reliable indicator of vesicle integrity and sample contamination ^37^. This ratio reflects the relative contribution of vesicular lipids and co-isolated soluble or proteinaceous material, with lower values corresponding to lipid-rich, vesicle-pure preparations, and higher ratios indicating protein contamination or vesicle disruption. According to the reference ranges established by Osteikoetxea et al., apoptotic bodies typically exhibit P/L ratios above 10, microvesicles between 1 and 10, and exosomes below 1. When compared with these values, the MPE-EVs obtained using the IA method (P/L ≈ 1.3 ± 0.2) correspond to highly pure small vesicles with minimal contamination by non-vesicular proteins. The NOR-derived MPE-EVs (P/L ≈ 12.8 ± 0.9) displayed intermediate purity, likely representing a mixed population of vesicles and soluble proteins, whereas the UC isolates (P/L ≈ 92.7 ± 4) showed a markedly elevated ratio indicative of substantial protein contamination and aggregation, consistent with the morphological findings. These results further support that the IA method yields MPE-EVs of superior purity, and highlight the relevance of reporting the P/L ratio as a complementary and quantitative parameter for evaluating EV preparation quality in both research and clinical settings.

In addition to the three methods tested in our study, several alternative techniques have been explored in previous studies. Size-exclusion chromatography (SEC) is simple, rapid, cost-effective, and yields relatively pure EV preparations, but it cannot effectively distinguish EVs from similarly sized particles or microbubbles ^38^. Density gradient ultracentrifugation (DGU), whether on continuous or discontinuous gradient, provides highly pure EVs but remains labor-intensive, requires specialized personnel and equipment, and is prone to lipoprotein contamination and EV aggregation ^38,39^. Another promising approach is ExoProK, which incorporates proteinase K pretreatment of PE samples to digest non-EV proteins such as albumin. This strategy improved EV purity and morphology, highlighting the importance of sample preprocessing ^40^ especially for downstream transcriptomic analysis.

Given that no single method can simultaneously optimize both yield and purity, combining complementary techniques has been suggested ^41–43^. However, widespread implementation of such multi-step workflows in clinical settings remains impractical due to complexity, cost, and time demands. For EV-based liquid biopsies to achieve clinical utility, isolation methods must be standardized, cost-effective, automated, and scalable, while remaining user-friendly and feasible in routine laboratory workflows ^44^. Commercial kits have been developed with these goals in mind, yet challenges persist due to low purity and co-isolation of non-EV macromolecules ^45^. One possible solution lies in coupling existing methods—such as IC or NOR—with SEC, ideally following sample preprocessing steps such as proteinase K treatment or albumin depletion to enhance purity if nucleic acids are molecules of interest ^40,46^. Alternative, non-enzymatic preprocessing steps should be considered when EV protein markers are the main targets, as enzymatic treatment may disrupt the EV corona and compromise vesicle stability, potentially leading to results that do not accurately reflect the true state of the analyzed sample.

This study has several strengths. Fresh MPE samples were collected and processed within one hour before freezing at –80 °C, minimizing sample degradation. The use of pooled samples reduced interindividual variability, and only one type of MPE was analyzed, avoiding confounding factors associated with heterogeneous effusion types (e.g., hemorrhagic or eosinophilic effusions). However, some limitations should be acknowledged. Due to lack of MPE samples we did not perform EV proteomic study, which could give us more precise information about enriched EV subpopulations.

Overall, methodological inconsistencies, lack of standardized sample preprocessing, and insufficient reporting of MPE type contribute to challenges in comparing findings across different studies. While it is recognized that different biological fluids require distinct EV isolation strategies, establishing uniform guidelines for collection, preprocessing, and isolation of MPE samples will be essential for advancing the field and enabling meaningful comparison of results across studies.

## Conclusion

This study highlights critical differences in the efficiency, purity, and aggregation of EVs isolated from MPE using three different EV isolation methods. IA demonstrated the best overall performance, whereas UC was least effective, and the NOR yielded intermediate results with the advantage of reduced aggregation. These findings confirm that UC and NOR by itself are not capable of optimizing both yield and purity, underscoring the potential value of combined or sequential techniques. IA method has promising results, but further investigation is needed. To advance pleural effusion–based liquid biopsy toward clinical utility, the development of robust, scalable, and user-friendly EV isolation workflows, coupled with harmonized guidelines for sample collection, preprocessing, and reporting, will be essential. Establishing such standards will facilitate reproducibility, enable meaningful cross-study comparisons, and accelerate the integration of EV–based biomarkers into precision oncology.

## Data accessibility

The raw data supporting the findings of this study are available from the corresponding author upon reasonable request.

## Acknowledgements and funding

This study was funded by the Horizon Europe EXPAND-EV (HORIZON-MSCA-2023-SE-01, Grant Agreement No. 101182851) and the Ministry of Science, Technological Development and Innovation of the Republic of Serbia (Agreement No. 451-03-136/2025-03/ 200043).

## Author Contributions

M.V. Conceptualization, Data Curation, Formal Analysis, Investigation, Methodology, Visualization, Writing-original draft, Writing-review and editing; L.F. Data Curation, Formal Analysis, Investigation, Methodology, Writing-review and editing; N.P. Investigation, Formal Analysis, Methodology, Writing-review and editing, Supervision; A.Z. Data Curation, Methodology, Investigation, Writing-review and editing; M.K. Conceptualization, Data Curation, Formal Analysis, Investigation, Writing-original draft, Writing-review and editing; M.T. Funding acquisition, Project Administration, Supervision, Writing-review and editing; R.J. Funding acquisition, Project Administration, Supervision, Writing-review and editing; T.S. Funding acquisition, Project Administration, Supervision, Writing-review and editing; M.P. Conceptualization, Funding acquisition, Investigation, Methodology, Data Curation, Formal Analysis, Project Administration, Supervision, Writing-original draft, Writing-review and editing; A.K. Project Administration, Conceptualization, Formal Analysis, Investigation, Methodology, Supervision, Writing-review and editing; S.S. Formal Analysis, Methodology, Writing-review and editing; M.C. Conceptualization, Funding acquisition, Investigation, Methodology, Project Administration, Supervision, Writing-original draft, Writing-review and editing. All authors have read and agreed to the published version of the manuscript.

## Ethics approval

All analyses from this study were approved by institutional Ethics Committees of the University Clinical Center of Serbia and the Institute for Oncology and Radiology of Serbia (approval no. 3780/2 from 07.11.2022. and 01-1/2024/965 from 29.3.2024.) and all patients signed an informed consent.

## Glossary

AFM: Atomic Force Microscopy
CTCs: circulating tumor cells
EBUS-FNA: Endobronchial Ultrasound-Guided Fine-Needle Aspiration
ELBS: European society for liquid biopsy
EVs: extracellular vesicles
IA: Immunoaffinity capture
ISEV: International Society for Extracellular Vesicles
LDCT: low-dose computed tomography
lncRNA: long non-coding RNA
miRNA: microRNA
MPE: malignant pleural effusion
MRI: magnetic resonance imaging
MVB: multivesicular bodies
NOR: NORGEN commercial kit
NSCLC: Non-small cell lung cancer
NTA: Nanoparticle Tracking Analysis
PBS: phosphate buffered saline
PE: pleural effusion
SEM: Scanning Electron Microscopy
TEM: Transmission Electron Microscopy
UC: Ultracentrifugation

## Footnotes

* https://www.isev.org/output

## References

1. Bray F, Laversanne M, Sung H, et al. Global cancer statistics 2022: GLOBOCAN estimates of incidence and mortality worldwide for 36 cancers in 185 countries. CA Cancer J Clin. 2024;74(3):229–263. doi:10.3322/caac.21834

2. Boukouris AE, Michaelidou K, Joosse SA, et al. A comprehensive overview of minimal residual disease in the management of early-stage and locally advanced non-small cell lung cancer. npj Precis Oncol. 2025;9(1). doi:10.1038/s41698-025-00984-9

3. Yang L, Dou Y, Sui Z, et al. Upregulated miRNA 182 5p expression in tumor tissue and peripheral blood samples from patients with non small cell lung cancer is associated with downregulated Caspase 2 expression. Exp Ther Med. Published online 2019:603–610. doi:10.3892/etm.2019.8074

4. Ganti AK, Klein AB, Cotarla I, Seal B CE. Update of Incidence, Prevalence, Survival, and Initial Treatment in Patients With Non–Small Cell Lung Cancer in the US. JAMA Oncol. 2021;7(12):1824–1832. doi:10.1001/jamaoncol.2021.4932

5. Liang W, Zhao Y, Huang W, et al. Non-invasive diagnosis of early-stage lung cancer using high-throughput targeted DNA methylation sequencing of circulating tumor DNA (ctDNA). Theranostics. 2019;9(7):2056–2070. doi:10.7150/thno.28119

6. Liu QX, Zhou D, Han TC, et al. A Noninvasive Multianalytical Approach for Lung Cancer Diagnosis of Patients with Pulmonary Nodules. Adv Sci. 2021;8(13):1–12. doi:10.1002/advs.202100104

7. Chen Y, Zitello E, Guo R, Deng Y. The function of LncRNAs and their role in the prediction, diagnosis, and prognosis of lung cancer. Clin Transl Med. 2021;11(4). doi:10.1002/ctm2.367

8. Cavic M, Kovacevic T, Zaric B, et al. Lung Cancer in Serbia. J Thorac Oncol. 2022;17(7):867–872. doi:10.1016/j.jtho.2022.04.010

9. Nikanjam M, Kato S, Kurzrock R. Liquid biopsy: current technology and clinical applications. J Hematol Oncol. 2022;15(1):1–14. doi:10.1186/s13045-022-01351-y

10. Wang K, Wang X, Pan Q, Zhao B. Liquid biopsy techniques and pancreatic cancer: diagnosis, monitoring, and evaluation. Mol Cancer. 2023;22(1):1–25. doi:10.1186/s12943-023-01870-3

11. Dasari A, Morris VK, Allegra CJ, et al. ctDNA applications and integration in colorectal cancer: an NCI Colon and Rectal–Anal Task Forces whitepaper. Nat Rev Clin Oncol. 2020;17(12):757–770. doi:10.1038/s41571-020-0392-0

12. Vukovic M, Tanic M, Damjanovic A, et al. EGFR mutation testing from pleural effusions of non-small cell lung cancer patients at the institute for oncology and radiology of Serbia. Transl Oncol. 2023;37(August):101772. doi:10.1016/j.tranon.2023.101772

13. Gonnelli F, Hassan W, Bonifazi M, et al. Malignant pleural effusion: current understanding and therapeutic approach. Respir Res. 2024;25(1):1–11. doi:10.1186/s12931-024-02684-7

14. Kaul V, McCracken DJ, Rahman NM, Epelbaum O. Contemporary approach to the diagnosis of malignant pleural effusion. Ann Am Thorac Soc. 2019;16(9):1099–1106. doi:10.1513/AnnalsATS.201902-189CME

15. Okamoto T, Iwata T, Mizobuchi T, et al. Pulmonary resection for lung cancer with malignant pleural disease first detected at thoracotomy. Eur J Cardio-thoracic Surg. 2012;41(1):25–30. doi:10.1016/j.ejcts.2011.04.010

16. Yao X, Liao B, Chen F, et al. Comparison of proteomic landscape of extracellular vesicles in pleural effusions isolated by three strategies. Front Bioeng Biotechnol. 2023;11(April):1–14. doi:10.3389/fbioe.2023.1108952

17. Sorolla MA, Sorolla A, Parisi E, Salud A, Porcel JM. Diving into the pleural fluid: Liquid biopsy for metastatic malignant pleural effusions. Cancers (Basel). 2021;13(11):1–25. doi:10.3390/cancers13112798

18. Guo L, He B. Extracellular vesicles and their diagnostic and prognostic potential in cancer. Transl Cancer Res. 2017;6(3):599–612. doi:10.21037/tcr.2017.06.32

19. Buzas EI. The roles of extracellular vesicles in the immune system. Nat Rev Immunol. 2023;23(4):236–250. doi:10.1038/s41577-022-00763-8

20. Mohammadi S, Yousefi F, Shabaninejad Z, et al. Exosomes and cancer: From oncogenic roles to therapeutic applications. IUBMB Life. 2020;72(4):724–748. doi:10.1002/iub.2182

21. Nawaz M, Fatima F, Vallabhaneni KC, et al. Extracellular Vesicles: Evolving Factors in Stem Cell Biology. Stem Cells Int. 2016;2016. doi:10.1155/2016/1073140

22. Valadi H, Ekström K, Bossios A, Sjöstrand M, Lee JJ, Lötvall JO. Exosome-mediated transfer of mRNAs and microRNAs is a novel mechanism of genetic exchange between cells. Nat Cell Biol. 2007;9(6):654–659. doi:10.1038/ncb1596

23. Kalluri R, McAndrews KM. The role of extracellular vesicles in cancer. Cell. 2023;186(8):1610–1626. doi:10.1016/j.cell.2023.03.010

24. Chee TM, O’Farrell HE, Lima LG, et al. Optimal isolation of extracellular vesicles from pleural fluid and profiling of their microRNA cargo. J Extracell Biol. 2023;2(10). doi:10.1002/jex2.119

25. Lucien F, Teske JJ, Gustafson D, et al. MIBlood-EV: Minimal information to enhance the quality and reproducibility of blood extracellular vesicle research. 2023;(November). doi:10.1002/jev2.12385

26. Sandau US, Ikezu T, Vella LJ, et al. Recommendations for reproducibility of cerebrospinal fluid extracellular vesicle studies. 2024;(June 2023). doi:10.1002/jev2.12397

27. Filipović L, Spasojević M, Prodanović R, et al. Affinity-based isolation of extracellular vesicles by means of single-domain antibodies bound to macroporous methacrylate-based copolymer. N Biotechnol. 2022;69(February):36–48. doi:10.1016/j.nbt.2022.03.001

28. Loveland P, Christie M, Hammerschlag G, Irving L, Steinfort D. Diagnostic yield of pleural fluid cytology in malignant effusions: an Australian tertiary centre experience. Intern Med J. 2018;48(11):1318–1324. doi:10.1111/imj.13991

29. Andre F, Schartz NEC, Movassagh M, et al. Malignant effusions and immunogenic tumour-derived exosomes. Lancet. 2002;360(9329):295–305. doi:10.1016/S0140-6736(02)09552-1

30. Zhu LR, Yuan RX, Xia X Bin, et al. Assessment of a panel of miRNAs in serum and pleural fluid for the differential diagnosis of malignant and benign pleural effusion. Cancer Biomarkers. 2022;33(1):71–82. doi:10.3233/CBM-210090

31. Tamiya H, Mitani A, Saito A, et al. Exosomal MicroRNA expression profiling in patients with lung adenocarcinoma-associated malignant pleural effusion. Anticancer Res. 2018;38(12):6707–6714. doi:10.21873/anticanres.13039

32. Shojaee S, Romano G, Sanchez TM, et al. Extracellular Vesicle MicroRNA in Malignant Pleural Effusion. Genes (Basel). 2022;13(11). doi:10.3390/genes13112159

33. An, M., Wu, J., Zhu, J., & Lubman DM. Comparison of an Optimized Ultracentrifugation Method versus Size-Exclusion Chromatography for Isolation of Exosomes from Human Serum. 2019;17(10):3599–3605. doi:10.1021/acs.jproteome.8b00479.The

34. Liu W zhao, Ma Z jun, Kang X wen. Current status and outlook of advances in exosome isolation. Anal Bioanal Chem. Published online 2022:7123–7141. doi:10.1007/s00216-022-04253-7

35. Gelibter S, Marostica G, Mandelli A, et al. The impact of storage on extracellular vesicles: A systematic study. J Extracell Vesicles. 2022;11(2). doi:10.1002/jev2.12162

36. Cavallaro, S., Hååg, P., Viktorsson, K., Krozer, A., Fogel, K., Lewensohn, R., … & Dev A. Nanoscale Advances Comparison and optimization of nanoscale extracellular vesicle imaging by scanning electron microscopy for accurate size-based profiling and morphological analysis. Nanoscale Adv. 2021;3(11):3053–3063. doi:10.1039/D0NA00948B

37. Osteikoetxea X, Balogh A, Szabó-taylor K, Németh A. Improved Characterization of EV Preparations Based on Protein to Lipid Ratio and Lipid Properties. PLoS One. 2015;10(3):1–16. doi:10.1371/journal.pone.0121184

38. Sidhom K, Obi PO, Saleem A. A review of exosomal isolation methods: Is size exclusion chromatography the best option? Int J Mol Sci. 2020;21(18):1–19. doi:10.3390/ijms21186466

39. Yuana Y, Levels J, Grootemaat A, Sturk A, Nieuwland R. Co-isolation of extracellular vesicles and high-density lipoproteins using density gradient ultracentrifugation. J Extracell Vesicles. 2014;3(1):1–5. doi:10.3402/jev.v3.23262

40. Antonopoulos D, Tsilioni I, Tsiara S, et al. Exoprok: A practical method for the isolation of small extracellular vesicles from pleural effusions. Methods Protoc. 2021;4(2):1–14. doi:10.3390/mps4020031

41. Koh YQ, Almughlliq FB, Vaswani K, Peiris HN, Mitchell MD. Exosome enrichment by ultracentrifugation and size exclusion chromatography. Front Biosci - Landmark. 2018;23(5):865–874. doi:10.2741/4621

42. Ryu, K. J., Lee, J. Y., Park, C., Cho, D., & Kim SJ. Isolation of Small Extracellular Vesicles From Human Serum Using a Combination of Ultracentrifugation With Polymer-Based Precipitation. Ann Lab Med. 2020;40(3):209–215. doi:10.3343/alm.2020.40.3.253

43. Zhang X, Borg EGF, Liaci AM, Vos HR, Stoorvogel W. A novel three step protocol to isolate extracellular vesicles from plasma or cell culture medium with both high yield and purity. J Extracell Vesicles. 2020;9(1). doi:10.1080/20013078.2020.1791450

44. Brennan K, Martin K, FitzGerald SP, et al. A comparison of methods for the isolation and separation of extracellular vesicles from protein and lipid particles in human serum. Sci Rep. 2020;10(1):1–13. doi:10.1038/s41598-020-57497-7

45. Patel GK, Khan MA, Zubair H, et al. Comparative analysis of exosome isolation methods using culture supernatant for optimum yield, purity and downstream applications. Sci Rep. 2019;9(1):1–10. doi:10.1038/s41598-019-41800-2

46. Reale A, Carmichael I, Xu R, et al. Human myeloma cell- and plasma-derived extracellular vesicles contribute to functional regulation of stromal cells. Proteomics. 2021;21(13-14):1–19. doi:10.1002/pmic.202000119

